# Correlation between serum 25(OH) vitamin D level and sleep quality in elderly patients with type 2 diabetes

**DOI:** 10.1101/2020.09.25.20201756

**Authors:** Hongyan Ma, Danni Fan, Xiaofei Li, Ying Qi, Yang Zhao, Lin Zhang

## Abstract

**Background:** Untreated sleep disorders have been linked with type 2 diabetes in previous evidence. This study was to investigate the correlation between serum 25(OH) vitamin D level and sleep quality in elderly patients with type 2 diabetes and reveal the therapeutic potential of 25(OH) vitamin D-based treatments to improve sleeping quality and its effect on diabetes management.

**Methods:** The sleep quality of 186 elderly patients with type 2 diabetes were assessed by questionaires of Pittsburgh sleep index (PSQI), with a cutoff point of PSQI • 7 defined as poor sleep. The measurement of serum 25(OH) vitamin D, fasting blood glucose, glycosylated hemoglobin, serum creatinine, total cholesterol, triglyceride, low-density lipoprotein cholesterol, and urinary albumin/creatinine ratio were collected. History of diabetic nephropathy, diabetic retinopathy, peripheral neuropathy, blood pressure, height, weight, hypoglycemic drug use, and the duration of diabetes also were recorded. Both univariate and multivariate logistic regression analysis was used to investigate the relationship of serum 25(OH) vitamin D and sleep quality in elderly patients with type 2 diabetes.

**Results:** The age of elderly patients with type 2 diabetes was 73.6±4.3 years., and duration of diabetes was 8.8 ± 3.0 years. In the study, inadequate quality of sleep was reported in 63% elderly patients with type 2 diabetes. This study revealed that lower level of 25(OH) vitamin D was significantly associated with poor sleep quality in type 2 diabetes patients(P < 0.05). Multivariate logistic regression analysis showed that 25(OH) vitamin D deficiency and the use of insulin were risk factors for of inadequate sleep quality in elderly patients with type 2 diabetes.

**Conclusions:** Elderly type 2 diabetic patients with 25(OH) vitamin D deficiency showed a high incidence of poor sleep quality, low-level of 25(OH) vitamin D was a risk factor for inadequate quality of sleep in elderly patients with type 2 diabetes.

## 1. Introduction

Type 2 diabetes is a chronic endocrine ailment, its prevalence rate continues to rise because of the change of lifestyle and is as high as 10% China and India^1^. Type 2 diabetes is estimated that around 60 million in Europe had type 2 diabetes by 2017 and would cause serious cardiovascular effects^2^. Type 2 diabetes and sleep disorders both are common health problems and affect each other. Persons with diabetes suffered from sleep disorders and took more sleeping medications ^3 4^. According to latest statistics, more than 30% of adults suffer from sleep disorders, and the incidence of sleep disorders in the elderly over 60 years old is up to 40%^5 6^. Type 2 diabetes has shown to be associated with higher incidence of sleep disorders. Sleep disorders might be caused by the disease itself or by complications of the disease, such as peripheral neuropathy and polyuria.

25(OH) vitamin D is essential for good bone health and deficiency or insufficiency have also been linked to other health concerns, such as diabetes, cardiovascular disease, hypertension, tumor and autoimmune diseases^7^. The prevalence of Vitamin D deficiency has increased in recent decades.^8 9^ In a recent population-based study of Asian adults, approximately 75% had suboptimal Vitamin D concentrations.^10^ The Endocrine Society Clinical Practice Guideline defines Vitamin D deficiency as 25(OH)D level <20 ng/mL and insufficiency as 21 to 29 ng/mL. ^11^ Some studies have linked deficient 25(OH) vitamin D to neuropsychological diseases such as sleep disorders^12-14^, which showed that 25(OH) vitamin D deficiency was associated with a higher risk of inadequate quality of sleep. Massa, Jennifer et al. found a significant trend that lower total serum 25(OH) vitamin D was associated with shorter sleep duration, and that men with the lowest total serum 25(OH) vitamin D(< 20.3 ng/mL) were twice as likely to have a shorter sleep duration than those with the highest total serum 25(OH) vitamin D (•40.06 ng/mL). Some researchers believed that the neural systems that control the opposing states of arousal and sleep are predominantly confined to the brainstem, hypothalamus and the thalamus.It has been shown that 25(OH) vitamin D target neurons are thought to be involved in sleep regulation^15^.

There was a statistical difference between 25(OH) vitamin D concentration in diabetic patients and the healthy subjects in the study by Mohammad Ali Bayani. et al. The results showed that the mean concentration of healthy women was higher than that 25(OH) vitamin D in diabetic women (27.03±10.28 ng/dl, 19.3±11.9 ng/dl, respectively) (P =0.0001) ^16^ Therfore, we may believe that 25(OH) vitamin D is important for diabetic women. In addition, a study was been shown that 25(OH) vitamin D affects sleep in people between the ages of 20 and 50. 25(OH) vitamin D supplementation can improve the sleep quality, reduce the sleep latency, prolong the sleep time, and improve the subjective sleep quality of patients with sleep disorders. However, there are still few studies on the effect of 25(OH) vitamin D on sleep quality in elderly patients with type 2 diabetes^17^.

This study was designed to investigate the relationship between serum 25(OH) vitamin D levels and sleep quality in elderly patients with type 2 diabetes in China and to fill in the literature gap on this topic.

## 2. Materials and Methods

### 2.1 Patients and definitions

A total of 186 inpatients were selected, who were admitted in the Department of Endocrinology and Geriatric Medicine of the First Affiliated Hospital of the University of Science and Technology of China (USTC) from January 2019 to December 2019. All patients were over 60 years old and diagnosed with type 2 diabetes according to the 1999 WHO Diabetes Diagnostic Criteria. Exclusion criteria: Patients with type 1 diabetes and other special types of diabetes; with type 2 diabetes-related acute complications; with thyroid and parathyroid function disorders; with cardiac insufficiency, hepatic and renal dysfunction, respiratory failure, malignant tumors, and other severe illness; with incorporative psychiatric illnesses, mental retardation and cognitive deficit; being treated with glucocorticoids, 25(OH) vitamin D, calcium and other drugs that may affect 25(OH) vitamin D levels in the past 3 months. Patient informed consents were obtained. All the data received Institutional Review Board (IRB) approval by the Ethics Committee.

### 2.2 Study design and Method

#### 2.2.1 Clinical data collection

The age, blood pressure, height, body weight, the duration of type 2 diabetes patients, medications (including oral hypoglycemic drugs and insulin) of all patients were recorded. Body mass index (BMI) was calculated according in kg/m ^2^. All patients were subjected for fundus examination to identify diabetic retinopathy after mydriasis, excluding other causes of retinopathy and physical examination and nerve conduction velocity (NCV) examination to identify diabetic peripheral neuropathy, excluding other neurological diseases and other causes of peripheral neuropathy. Venous fasting blood was sampled including fasting blood glucose (FBG), glycosylated hemoglobin (HbA1c), serum creatinine (Cr), total cholesterol (TC), triacylglycerol (TG) and low-density lipoprotein (LDL-C) were measured. Urine albumin was assayed by immunoturbidimetry, and urine creatinine concentration was determined by benedict-behre creatinine colorimetry urine albumin/creatinine ratio (UACR) was calculated, and diabetic nephropathy was defined by an UACR of •30mg/g.

#### 2.2.2 Sleep quality assessment

The Pittsburgh Sleep Quality Index (PSQI) was originally developed in 1989 to measure sleep quality for psychiatric research as well as clinical practice^18^. The index has since been used to assess sleep quality in patients suffering T2DM^19^. The PSQI scale was used to evaluate the sleep status of all subjects in the past 4 weeks. The questionnaire included 19 self-assessments and 5 other assessment items. Higher scores indicated worse sleep quality. The investigator made use of questionnaire adjustment method. The adjustment tables were independently completed by the study subjects. Those who could not be accomplished independently due to visual impairment, low cultural level, etc., were asked by the investigator according to the content of the scale and the answer results were truthfully recorded. Poor sleep quality was defined by a total PSQI score ^20^ of >7.

### 2.3 Statistical analyses

Normal distribution was determined by the Kolmogorove-Smirnov test, and the data were expressed as mean ± standard deviation. Group comparison was performed by t test. The data that did not meet the normal distribution was compared by the Mann-Whitney U test. Count data were reported in rates, and comparisons between groups were performed using the •2 test. Univariate logistic regression analysis was used to screen variables for selected research factors. Univariate analysis was performed with the presence of sleep disturbance as the dependent variable and gender, age, body mass index (BMI), duration of diabetes, systolic blood pressure, diastolic blood pressure, FBG, HbA1c, TG, TC, LDL-C, Cr, 25(OH) vitamin D, diabetic nephropathy, diabetic peripheral neuropathy, diabetic retinopathy, and insulin use as the independent variables. Multivariate logistic regression analysis was used to explore the influencing factors of sleep disorder in elderly patients with type 2 diabetes. All tests were two-tailed test. A P value of <0.05 indicated statistically significant. SPSS software version 22.0 (SPSS Inc. Chicago, IL, USA,) was used for statistical analysis of the data.

## 3. Results

### 3.1 Baseline characteristics of two groups accroding to sleep quality in the elderly patients with type 2 diabetes

186 type 2 diabetes elderly patients were recruited in the study with average age (73.6±4.3) years and the average duration of diabetes (8.8±3.0) years. There were 68 cases with diabetic nephropathy, 62 cases with diabetic retinopathy, and 50 cases with diabetic peripheral neuropathy; 100 cases were treated with insulin. According to the total PSQI score, 186 elderly patients with type 2 diabetes were divided into a normal sleep group (PSQI •7) and an inadequate sleep quality group (total PSQI score > 7). 117 cases presented with poor sleep quality, accounting for 63% of all subjects under study. Compared with the normal sleep group, the inadequate sleep quality group had lower 25(OH) vitamin D level, higher HbA1c level, and a higher incidence of diabetic nephropathy and more insulin use (P<0.05). The differences in gender, age, BMI, duration of diabetes, systolic blood pressure, diastolic blood pressure, FBG, TG, TC, LDL-C, and Cr levels in the two groups were not statistically significant (all P>0.05). The results were showed in **Table 1**.

**Table 1.** Baseline characteristics of two groups accroding to sleep quality in the elderly patients with type 2 diabetes

|  | Normal sleep group<br>( n=69 ) | Inadequate sleep group<br>( n=117 ) | t ( •<sup>2</sup> ) | P |
| --- | --- | --- | --- | --- |
| Sex (male/female) | 46/23 | 76/41 | 0.056 | 0.813 |
| Age (years) | 73.0±4.1 | 73.9±4.3 | -1.295 | 0.197 |
| BMI (kg/m<sup>2</sup>) | 21.5±2.9 | 21.3±3.4 | 0.425 | 0.672 |
| Duration of diabetes (years) | 8.6±2.8 | 8.9±3.1 | -0.623 | 0.534 |
| Systolic pressure (mmHg) | 129.5±8.2 | 131.9±8.2 | -1.931 | 0.055 |
| Diastolic pressure (mmHg) | 74.1±7.8 | 76.0±7.9 | -1.514 | 0.132 |
| FBG (mmol/L) | 7.24±1.9 | 7.52±2.35 | -0.855 | 0.393 |
| HbA1c (%) | 6.43±2.6 | 7.35±2.69 | -2.274 | 0.024 |
| TG (mmol/L) | 1.53±0.7 | 1.67±0.67 | -1.308 | 0.192 |
| TC (mmol/L) | 4.54±1.3 | 4.92±1.68 | -1.645 | 0.102 |
| LDL-C (mmol/L) | 3.87±0.9 | 4.14±0.96 | -1.824 | 0.07 |
| Cr (•mol/L) | 88.77±23.1 | 92.42±29.2 | -0.888 | 0.376 |
| 25(OH) vitamin D (ng/mL) | 20.674±2.6 | 13.131±2.9 | 18.004 | <0.01 |
| Diabetic nephropathy (%) | 18 (26.1) | 50 (42.7) | 5.187 | 0.023 |
| Diabetic peripheral neuropathy (%) | 16 (23.2) | 34 (29.1) | 0.761 | 0.383 |
| Diabetic retinopathy (%) | 20 (29.0) | 42 (35.9) | 0.933 | 0.334 |
| Insulin use (%) | 25 (36.2) | 75 (64.1) | 13.563 | <0.01 |

### 3.2 *Comparison of sleep quality in different* 25(OH) vitamin D *level groups in elderly patients with type 2 diabetes*

There were 141 cases presented with 25(OH) vitamin D deficiency, accounting for 76% of the 186 elderly patients with type 2 diabetes recruited in this study. The average PSQI total score of the 25(OH) vitamin D-deficient group was higher than that of the 25(OH) vitamin D -nondeficient group, and the difference was statistically significant (P <0.05); The scores of 25(OH) vitamin D deficient group were higher than those of normal 25(OH) vitamin D group (P <0.05) in 7 dimensions of subjective sleep quality, sleep latency, sleep duration, habitual sleep efficiency, sleep disturbances, use of sleeping medication and daytime dysfunction (**Table 2**).

**Table 2.** Comparison of sleep quality in two groups, which were divided based on vitamin D level in the elderly patients with type 2 diabetes

|  | 25(OH) vitamin D<br>-nondeficient group<br>(n=45) | 25(OH) vitamin D<br>-deficient group<br>(n=141) | Z ( • 2 ) | P |
| --- | --- | --- | --- | --- |
| Sex (male/female) | 31/14 | 91/50 | 0.286 | 0.593 |
| PSQI total score | 6.69±1.4 | 10.6±1.9 | -9.190 | <0.01 |
| Subjective sleep quality | 1.47±0.84 | 1.81±0.88 | -2.329 | 0.02 |
| Sleep latency | 0.96±0.77 | 1.54±0.94 | -3.664 | <0.01 |
| Sleep duration | 0.98±0.15 | 1.28±0.83 | -2.565 | <0.01 |
| Habitual sleep efficiency | 0.91±0.56 | 1.46±0.64 | -4.903 | <0.01 |
| Sleep disturbances | 0.89±0.32 | 1.65±0.49 | -7.856 | <0.01 |
| Use of sleeping medication | 0.38±0.49 | 0.70±0.46 | -3.900 | <0.01 |
| Daytime dysfunction | 0.86±0.34 | 2.15±0.90 | -7.995 | <0.01 |

### 3.3 Logistic regression analysis of risk factors for sleep disorders in elderly patients with type 2 diabetes

The results showed diabetic nephropathy, HbA1c, 25(OH) vitamin D, use of insulin were related to the occurrence of inadequate quality of sleep in patients. Multivariate logistic regression analysis of the above indicators suggested that 25(OH) vitamin D deficiency and the use of insulin were risk factors for of inadequate sleep quality in elderly patients with type 2 diabetes. (**Table 3**)

**Table 3.** logistic analysis of sleep quality risk factors in elderly patients with type 2 diabetes

| Risk factors | Exp(B) | 95% CI | P |
| --- | --- | --- | --- |
| 25(OH) vitamin D | 0.21 | 0.11- 0.41 | <0.01 |
| Use of Insulin * | 28.68 | 2.54-323.54 | <0.01 |

\* Use of insulin , no = 0 , yes = 1

## 4. Discussion

In this study, we revealed the correlation between serum 25(OH) vitamin D deficiency and sleep quality in elderly patients with type 2 diabetes.

Sleep plays an important role in human health, and sleep disorders is closely related to diabetes^21^. In recent years, sleep disorders have been taken into account as an important risk factor for the development of type 2 diabetes in the elderly. Sleep disorders and depression are more prevalent in people with type 2 diabetes. Van Dijk et al. reported in the study that the probability of increased nocturnal enuresis, pain, dysthermesthesia, and habitual snoring in diabetic populations were significantly increased ^22^. Epidemiological studies found that the incidence of sleep disorders in people with type 2 diabetes was 45% to 55%^23 24^. This study showed the prevalence of inadequate sleep quality, which is only a subset of all various sleep disorders, was high up to 63% in elderly patients with type 2 diabetes. This indicated the incidence of sleep disorders would be significantly higher in elderly patients among all type 2 diabetic patients^5^.

Inadequate sleep quality could cause sympathetic nerve excitement, increase the secretion of antagonist hormone of insulin such as nighttime growth hormone, glucagon, catecholamine, and cortisol, disturb glucose homeostasis; and increase insulin resistance^25^. One has shown One study also showed that long-term sleep deprivation could reduce insulin secretion in the body leading to increased blood glucose levels in patients with diabetes^26^. This study also found that HbA1c levels in diabetic patients with sleep disorders were higher than those in the normal sleep group, indicating that blood glucose in the sleep disorder group was more difficult to control.

Since serum 25(OH) vitamin D concentration is affected by many factors, including sun exposure, lifestyle, and skin tone, etc. 25(OH) vitamin D deficiency has become a worldwide problem and the prevalence is rising^27^. Bayani et al. found that compared with healthy controls, serum 25(OH) vitamin D levels were significantly lower in patients with type 2 diabetes, and the decline was more pronounced in women^28^. This study also showed that 25(OH) vitamin D deficiency was prevalent in elderly people with type 2 diabetes, accounting for 76% of the study population. The potential causes of low 25(OH) vitamin D in elderly patients could be less outdoor activities, shorter sunshine hours, and reduced gastrointestinal digestion and absorption in elderly patients.

The data in this study show that in elderly patients with type 2 diabetes, the serum 25(OH) vitamin D level in the sleep disorders group was lower than that in the normal sleep group, and the insulin use rate was higher than in the normal sleep group. The average total PSQI score of the 25(OH) vitamin D -deficient group was higher than those of the normal vitamin D group, and the 25(OH) vitamin D -deficient group scored higher in 7 dimensions of subjective sleep quality, sleep latency, sleep duration, habitual sleep efficiency, sleep disturbances, use of sleeping medication and daytime dysfunction. Further logistic regression analysis showed that low level of 25(OH) vitamin D and more insulin use were associated with sleep disturbance in elderly patients with type 2 diabetes. Logistic regression analysis of risk factors for sleep disorders in elderly patients with type 2 diabetes demonstrated that the low-level serum 25(OH) vitamin D was an important risk factor for inadequate quality of sleep in elderly patients with type 2 diabetes.

Knhanes result showed that lower serum 25(OH) vitamin D levels were linked to a significantly higher risk of short sleep duration^29 30^. McCarty et al. found that patients with 25(OH) vitamin D deficiency scored lower on the Epworth sleepiness scale (ESSs)^31^. Another study measured objective sleep duration and sleep effectiveness of men over the age of 65 and found that lower serum 25(OH) vitamin D levels were associated with shorter sleep duration and poorer sleep effectiveness^32^. These results mentioned above were consistent with our data.

So far, the mechanisms for a link between sleep duration and 25(OH) vitamin D were not clearly understood. Vitamin D receptors (VDR) were shown to be present in nearly all tissues of the body, including many parts of the human brain, such as the hypothalamus, prefrontal cortex, midbrain central gray, and substantia nigra, etc., which all play an important role in sleep regulation^32-34^.

The classical clinical; presentation of Vitamin D deficiency is bone patient. Increased pain could be associated with sleep disturbances^35^. Epidemiologic evidence for impact of 25(OH) vitamin D supplements on sleep quality was emerging. One study showed that people’s pain levels, sleep quality, and various aspects of Quality of Life (QoL) can be significantly improved after standardized vitamin D supplementation^36^. Another double-blind clinical trial showed improvements and increases in sleep quality and duration in patients with sleep disorders after eight weeks of continuous supplementation with 25(OH) vitamin D (50,000 IU/ 2 weeks)^17^.

As far as we know, this was the first study to analyze the relationship between serum 25(OH) vitamin D concentration and sleep quality in elderly patients with type 2 diabetes. The significance of this study is that the concentration of 25(OH) vitamin D can be used to identify sleep disorders in elderly patients with type 2 diabetes. These findings have positive implications for the prevention and treatment of inadequate sleep quality in elderly patients with type 2 diabetes. One limitation of this study was that the dietary habits and psychological state of patients were not assessed, which, may also be related to sleep quality. Further, because of the character of the cross-sectional study, we could not conclude the causal relationship among 25(OH) vitamin D and sleep quality. Therefore, further studies with larger scale cohort studies and well-designed randomized controlled trials would be needed to verify this relationship and reveal possible mechanisms.

## 5 Conclusion

In conclusion, this study confirms that 25(OH) vitamin D deficiency was more common in elderly type 2 diabetic patients. There was a clear correlation between vitamin D deficiency and sleep quality in this population. This initial work warrants further larger scale studies of ethnic 25(OH) vitamin D variations and potential causal interrelationship between sleep quality, insulin usage and 25(OH) vitamin D deficiency state among elderly type 2 diabetic patients. Further studies are required to investigate possible biological mechanisms of the association between 25(OH) vitamin D deficiency, insulin usage and sleep quality, and to discover whether effective interventions for 25(OH) vitamin D supplementation could improve sleeping quality and effectiveness of insulin treatment in decrease the risk of geriatric syndromes in elders with type 2 diabetes.

## Data Availability

All the data was approved for use for this publication by First Affiliated Hospital of University of Science and Technology of China.

## Author Contributions

Conceptualization, Hongyan Ma, Lin Zhang; Methodology, Hongyan Ma, Lin Zhang; Writing – original draft, Hongyan Ma, Dani Fan, Lin Zhang; Writing – review & editing, Hongyan Ma, Dani Fan, Xiaofei Li,Yang Zhao, Lin Zhang.All authors have read and agreed to the published version of the manuscript.

## Acknowledgments

This study was supported by the First Affiliated Hospital of University of Science and Technology of China for data access.

## Ethical approval

Informed consents of patients were obtained for diagnosis and treatment, and the study protocol was approved by the First Affiliated Hospital of University of Science and Technology of China. All the data received Institutional Review Board (IRB) approval by the Ethics Committee.

## References

1. Gwatidzo SD, Stewart Williams J. Diabetes mellitus medication use and catastrophic healthcare expenditure among adults aged 50+ years in China and India: results from the WHO study on global AGEing and adult health (SAGE). BMC Geriatrics 2017;17(1) doi:10.1186/s12877-016-0408-x

2. Cho NH, Shaw JE, Karuranga S, et al. IDF Diabetes Atlas: Global estimates of diabetes prevalence for 2017 and projections for 2045. Diabetes Research & Clinical Practice 2018:271.

3. Skomro RP, Ludwig S, Salamon E, et al. Sleep complaints and restless legs syndrome in adult type 2 diabetics. Sleep Medicine 2001;2(5):417–22.

4. Association AD. 3. Comprehensive Medical Evaluation and Assessment of Comorbidities: Standards of Medical Care in Diabetes—2018. Diabetes Care 2018;41(Suppl 1):S28.

5. Vittoria E, Sisti D, Pascucci P, et al. [Prevalence of Obstructive Sleep Apnea Syndrome among holders of a category B driver’s license and among professionals in the Province of Pesaro-Urbino (Italy)]. Igiene E Sanita Pubblica 2017;73(6):605.

6. Owens, J. Insufficient Sleep in Adolescents and Young Adults: An Update on Causes and Consequences. Pediatrics 2014;134(3):e921.

7. Mayte M, Estrella CC, Isabel M, et al. Vitamin D: Effect on Haematopoiesis and Immune System and Clinical Applications. International Journal of Molecular Sciences 2018;19(9):2663-.

8. Holick MF, Chen TC. Vitamin D deficiency: a worldwide problem with health consequences. The American journal of clinical nutrition 2008;87(4):1080S–6S. doi:10.1093/ajcn/87.4.1080S [published Online First: 2008/04/11]

9. Hilger J, Friedel A, Herr R, et al. A systematic review of vitamin D status in populations worldwide. Br J Nutr 2014;111(1):23–45. doi:10.1017/S0007114513001840 [published Online First: 2013/08/13]

10. Man RE, Li LJ, Cheng CY, et al. Prevalence and Determinants of Suboptimal Vitamin D Levels in a Multiethnic Asian Population. Nutrients 2017;9(3) doi:10.3390/nu9030313 [published Online First: 2017/03/23]

11. Holick MF, Binkley NC, Bischoff-Ferrari HA, et al. Evaluation, treatment, and prevention of vitamin D deficiency: an Endocrine Society clinical practice guideline. J Clin Endocrinol Metab 2011;96(7):1911–30. doi:10.1210/jc.2011-0385 [published Online First: 2011/06/08]

12. Ewelina Łukaszyk, Katarzyna, et al. Cognitive Functioning of Geriatric Patients: Is Hypovitaminosis D the Next Marker of Cognitive Dysfunction and Dementia? Nutrients 2018

13. Hiller AL, Murchison CF, Lobb BM, et al. A randomized, controlled pilot study of the effects of vitamin D supplementation on balance in Parkinson’s disease: Does age matter? PLoS ONE 2018;13(9)

14. Pierrot-Deseilligny C, Souberbielle JC. Vitamin D and multiple sclerosis: An update. Multiple Sclerosis & Related Disorders 2017;14:35.

15. Gominak SC, Stumpf WE. The world epidemic of sleep disorders is linked to vitamin D deficiency. Medical Hypotheses 2012;79(2):132–35. doi:10.1016/j.mehy.2012.03.031

16. MA Bayani.,. Ra, BB, et al. Status of Vitamin-D in diabetic patients. 2014;5(1):3. [published Online First: 42]

17. Majid MS, Ahmad HS, Bizhan H, et al. The effect of vitamin D supplement on the score and quality of sleep in 20-50 year-old people with sleep disorders compared with control group. Nutr Neurosci 2018;21(7):511–19. doi:10.1080/1028415X.2017.1317395

18. Buysse DJ, Reynolds CF, 3rd, Monk TH, et al. The Pittsburgh Sleep Quality Index: a new instrument for psychiatric practice and research. Psychiatry research 1989;28(2):193–213. doi:10.1016/0165-1781(89)90047-4 [published Online First: 1989/05/01]

19. Telford O, Diamantidis CJ, Bosworth HB, et al. The relationship between Pittsburgh Sleep Quality Index subscales and diabetes control. Chronic Illn 2019;15(3):210–19. doi:10.1177/1742395318759587 [published Online First: 2018/02/23]

20. Liu HX, Lin J, Lin XH, et al. Quality of sleep and health-related quality of life in renal transplant recipients. International Journal of Clinical & Experimental Medicine 2015;8(9):16191–98.

21. Buxton OM, Marcelli E. Short and long sleep are positively associated with obesity, diabetes, hypertension, and cardiovascular disease among adults in the United States. Social ence & Medicine 2010;71(5):1027–36.

22. Dijk MV, Donga E, Dijk JGV, et al. Disturbed subjective sleep characteristics in adult patients with long-standing Type 1 diabetes mellitus. Diabetologia 2011;54(8):1967–76.

23. Luyster FS, Dunbar-Jacob J. Sleep Quality and Quality of Life in Adults With Type 2 Diabetes. Diabetes Educator 2011;37(3):347–55.

24. Amarabalan Rajendran, Shruthi, et al. Prevalence and correlates of disordered sleep in southeast asian indians with type 2 diabetes. Diabetes & metabolism journal 2012

25. P CF Lanfranco DE, Pasquale S, et al. Quantity and quality of sleep and incidence of type 2 diabetes: a systematic review and meta-analysis.[J]. Diabetes care,2010,33(2). Diabetes care 2010;33(2) doi:10.2337/dc09-1124

26. Darukhanavala A, Booth JN, Bromley L, et al. Changes in Insulin Secretion and Action in Adults With Familial Risk for Type 2 Diabetes Who Curtail Their Sleep. Diabetes Care 2011;34(10):2259–64.

27. Norman AW, Bouillon R. Vitamin D nutritional policy needs a vision for the future. Experimental Biology & Medicine 2010;235(9):1034–45.

28. Bayani MA, Akbari R, Banasaz B, et al. Status of Vitamin-D in diabetic patients. Caspian J Intern Med 2014;5(1):3. [published Online First: 42]

29. Lee HJ, Lee HD, Hwang HS, et al. Associations between Serum Vitamin D Level and Mean Hours of Sleep: 2011Korea National Health and Nutrition Examination Survey. Metabolism-Clinical and Experimental 2014;7(8):6346–61.

30. Kim JH, Chang JH, Kim DY, et al. Association between self-reported sleep duration and serum vitamin D level in elderly Korean adults. Journal of the American Geriatrics Society 2014;62(12):2327–32.

31. Mccarty DE, Reddy A, Keigley Q, et al. Vitamin D, Race, and Excessive Daytime Sleepiness. Journal of Clinical Sleep Medicine Jcsm Official Publication of the American Academy of Sleep Medicine 2012;8(6):693–7.

32. Massa J, Stone KL, Wei EK, et al. Vitamin D and Actigraphic Sleep Outcomes in Older Community-Dwelling Men: The MrOS Sleep Study. Sleep 2015;38(2):251–57. doi:10.5665/sleep.4408

33. A EG, B WB, A MM, et al. New clues about vitamin D functions in the nervous system. Trends in Endocrinology & Metabolism 2002;13(3):100–05.

34. Musiol IM, Stumpf WE, Bidmon HJ, et al. Vitamin D nuclear binding to neurons of the septal, substriatal and amygdaloid area in the Siberian hamster (Phodopus sungorus) brain. Neuroence 1992;48(4):841–48.

35. Monika H, Elsa S, Mullington JM. Elevated inflammatory markers in response to prolonged sleep restriction are associated with increased pain experience in healthy volunteers. Sleep (9):1145.

36. Huang W, Shah S, Long Q, et al. Improvement of Pain, Sleep, and Quality of Life in Chronic Pain Patients With Vitamin D Supplementation. Clinical Journal of Pain 2013;29(4):341–47.

